# The uncoupling of all-cause excess mortality from Covid-19 cases and associated hospitalizations in late winter and spring of 2022 in a highly vaccinated state

**DOI:** 10.1101/2022.07.07.22277315

**Authors:** Jeremy Samuel Faust, Benjamin Renton, Alexander Junxiang Chen, Chengan Du, Chenxue Liang, Shu-Xia Li, Zhenqiu Lin, Harlan M. Krumholz

**Author notes:** (corresponding author) Jeremy Samuel Faust, MD MS Brigham and Women’s Hospital, Harvard Medical School.

## Abstract

**Introduction:** Since March 2020, all-cause excess mortality—the number of all-cause deaths exceeding the baseline number of expected deaths—has been observed in waves coinciding with Covid-19 outbreaks in the United States. We recently described high levels of excess mortality in Massachusetts during the initial 8-week Omicron wave. However, whether excess mortality continued after that period—during which an outbreak of Omicron subvariants occurred—is unknown.

**Methods:** We applied seasonal autoregressive integrated moving averages to five years of pre-pandemic data provided by the Massachusetts Registry of Vital Records and Statistics (MRVRS) to project the weekly populations and expected deaths for the pandemic period. Observed deaths during the pandemic were also provided by MRVRS and are >99% complete for all study weeks.

**Results:** During the 18-week Omicron subvariant period (the week ending February 27, 2022, through June 26, 2022) the incidence of all-cause excess mortality was 0.1 per 100,000-person weeks, corresponding to 148 excess deaths (95%. CI -907 to 1153), representing a 97.1% decrease from the initial Omicron period (during which all-cause excess mortality was 4.0 per 100,000-person-weeks), and a 91.9% reduction from the Delta and Delta-Omicron transition period (during which all-cause excess mortality was 1.5 per 100,000-person-weeks), despite >226,000 reported new Covid-19 cases during the subvariant/spring period. However, Covid-19-associated hospitalizations were observed during the subvariant/spring 2022 period.

**Conclusion:** In a highly vaccinated state with a recent wave of SARS-CoV-2, all-cause excess mortality was uncoupled from new case counts, indicating the possibility of temporary protection from the most severe outcomes related to Covid-19 among high-risk individuals. However, given the possibility of waning immunity and the emerging of new variants, continued monitoring is warranted.

## Main Body

Since March 2020, excess mortality—the number of all-cause deaths exceeding the baseline number of expected deaths—has been observed in waves coinciding with Covid-19 outbreaks in the United States. However, since February, the reported number of Covid-19-associated deaths decreased, despite a notable spring wave of cases primarily due to Omicron subvariants (BA.2, BA.2.12.1, BA.4, BA.5).^1^ Until now, it has been unknown whether the spring 2022 Covid-19 wave in Massachusetts was associated with all-cause excess mortality.

Accordingly, we assembled population data (2014-2019) and weekly mortality data (January 2015-February 2020) provided by the Massachusetts Registry of Vital Records and Statistics (MRVRS) and applied seasonal autoregressive integrated moving averages to project the weekly number of expected deaths for the state for the pandemic period (week ending February 9, 2020– June 26, 2022). We summed age-specific mortality to create state-level estimates and additionally corrected for the lower-than-expected state population owing to cumulative excess mortality recorded during the pandemic.^2,3^ Weekly observed deaths provided by the MRVRS are >99% complete for all study weeks. Case, wastewater, and hospitalization data were accessed from publicly available data.^4^ Analyses were conducted with R (v.4.1.2). The MRVRS deemed the study exempt from institutional review board review.

In the 18-week period since BA.2, BA.2.12.1, BA.4, and BA.5 became prevalent (week ending February 27, 2022, Supplemental Figure 1), there have been 0.1 excess deaths per 100,000 person-weeks, corresponding to 148 excess deaths (95% CI -907–1153) (Figure, Panel A-B), despite at least 226,857 newly recorded cases, as evidenced by corresponding substantial spikes in SARS-CoV-2 wastewater levels and changes in testing volume (Supplemental Figure 2-3). This corresponds to a 97.1% reduction in excess mortality compared to the 8-week initial Omicron (B.1.1.529) wave, during which excess mortality was 4.0 per 100,000 person-weeks (2,239 excess deaths; 95% CI 1,746–2,733) and a 91.9% reduction in excess mortality compared to the combined 26-week Delta (B.1.617.2) and Delta-to-Omicron transition periods, during which excess mortality was 1.5 per 100,000 person-weeks (2,643 excess deaths; 95% CI 1,192– 4,094). However, new Covid-19-associated hospitalizations continued to occur during this period (Figure, Panel C), either reflecting background community prevalence, or that new Covid-19 cases exacerbated chronic medical illnesses amply to require emergency care, but not to cause proximate mortality.

**Figure.**
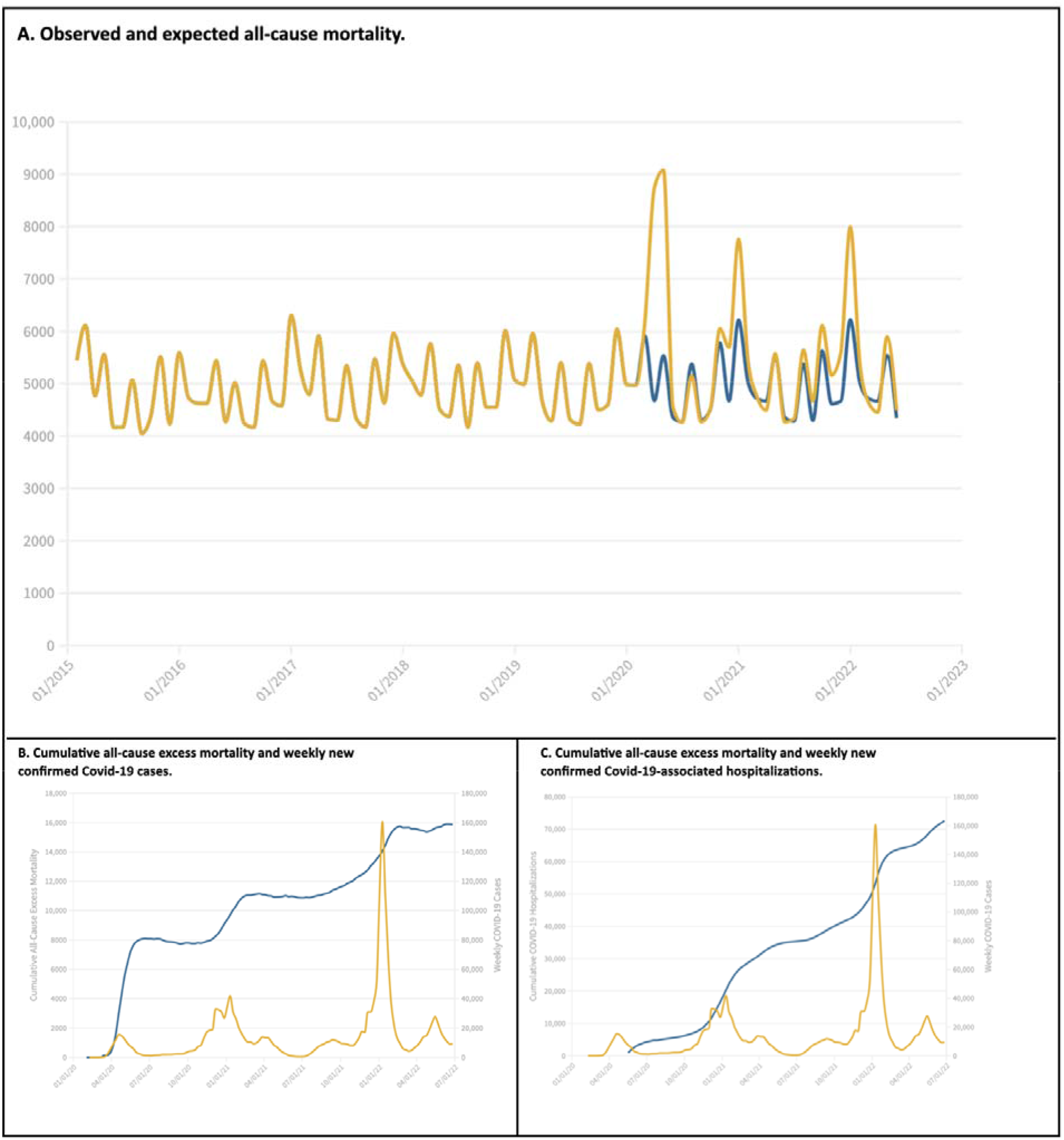
All-cause excess mortality, Covid-19-associated hospitalizations, and confirmed Covid-19 cases in Massachusetts. <u>Panel A</u> shows monthly number of observed deaths from February 2015 through June 2022 (yellow line) and the monthly number of expected deaths from February 2020 through June 2022 (blue line). The area between the orange and blue lines starting February 2020 correspond to all-cause excess mortality during the Covid-19 pandemic period. <u>Panel</u> B shows the cumulative all-cause excess mortality (blue line) plotted against weekly confirmed new Covid-19 cases (yellow line), Massachusetts, February 2020 through June 2022^5^. Note: hospitalization data did not become available until May, 2020. <u>Panel</u> C shows the cumulative Covid-19-associated hospitalizations (blue line) plotted against weekly confirmed new Covid-19 cases (yellow line), Massachusetts, February 2020 through June 2022.^5^

Since the initial Covid-19 outbreak in Massachusetts, there have been periods without excess mortality, corresponding to times of low prevalence (Figure, Panel B). However, we also have observed two substantial outbreaks not accompanied by excess mortality. The first instance, (late February-June, 2021) corresponded to the phased vaccine rollout period, during which the mean age of newly infected persons dropped precipitously, and prevalence among residents ages >60 years was low (Supplemental Appendix Figure 4), likely temporarily reflecting exceptionally high vaccine-conferred protection against SARS-CoV-2 infection among the vaccine-eligible population. The second instance occurred late February-June, 2022). Unlike February-June 2021, the mean age of newly infected persons did not fall during the corresponding 2022 period, and in fact rose.

The uncoupling of excess mortality and new Covid-19 cases, in the absence of decreases in the mean age of infected individuals (Supplemental Figure 4), suggests that in our highly vaccinated state, current levels of immunity are considerable, leaving many, if not most, high-risk individuals with substantial protection against the most feared outcomes of SARS-CoV-2 infection. However, given newly emerging variants and the unknown duration of protection, further monitoring is warranted.

## Data Availability

All data produced in the present study are available upon reasonable request to the authors

## Additional Contribution

We thank the Registry of Vital Records and Statistics, Office of Population Health, Massachusetts Department of Public Health, for assistance with data acquisition.

## Supplementary Appendix

This appendix has been provided by the authors to give readers additional information about the work.

## Supplemental figure legends

**Supplemental Figure 1.**
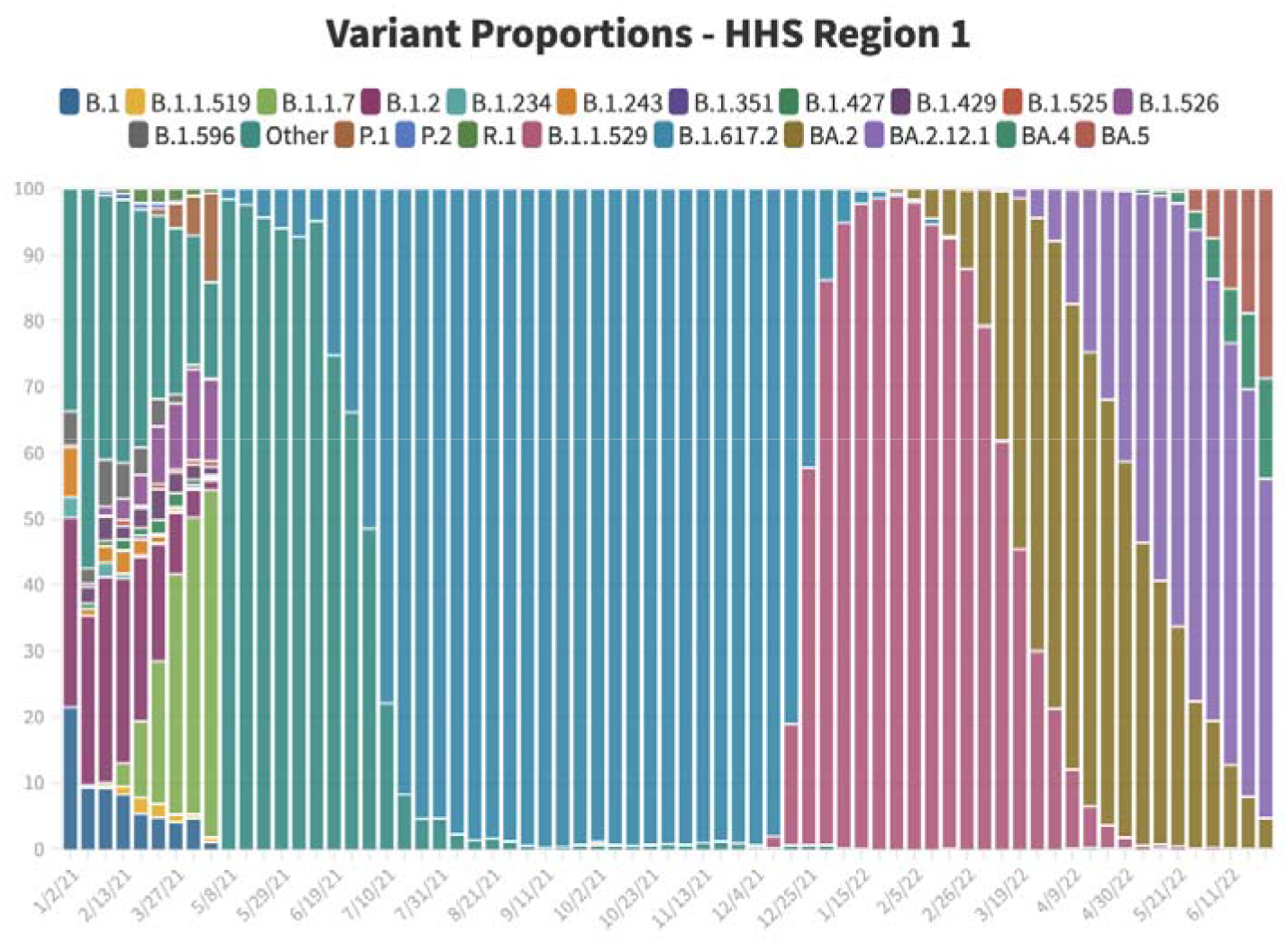
SARS-CoV-2 variant proportions for HHS Region 1 (Connecticut, Maine, Massachusetts, New Hampshire, Rhode Island, Vermont), January 2021 through early June 2022.^1^

**Supplemental Figure 2A.**
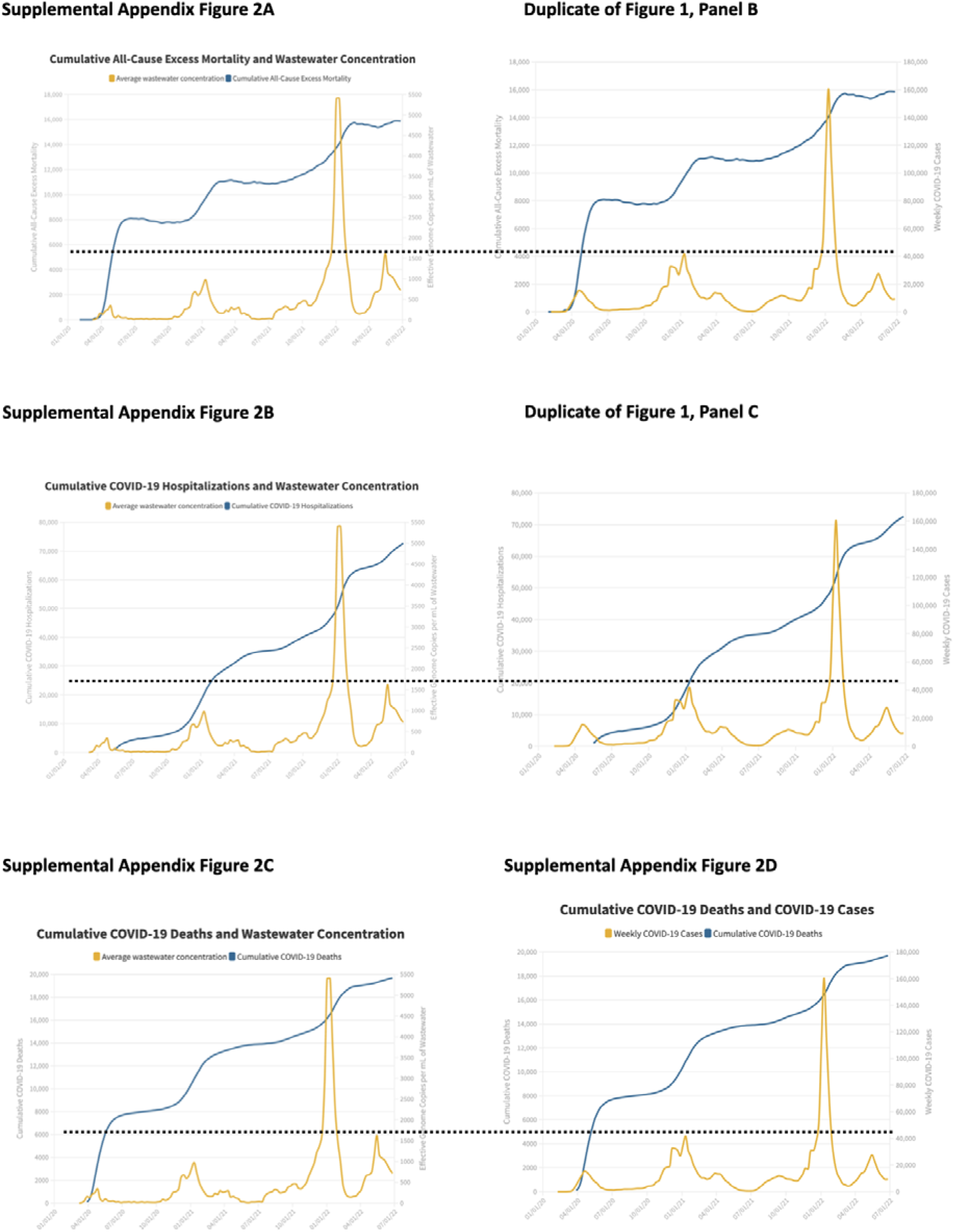
shows the cumulative all-cause excess mortality (blue line) plotted against weekly wastewater SARS-CoV-2 RNA copies per mL mean values (yellow line) from counties for which there were available data in a given week (Barnstable, Berkshire, Bristol, Essex, Franklin, Hampshire, Middlesex, Nantucket, Plymouth, Suffolk, Worcester), Massachusetts, February 2020 through June 2022 in Massachusetts. ^5^ Figure 1, Panel B is shown for comparison between documented cases and wastewater concentrations. Supplemental Figure 2B shows the cumulative Covid-19-associated hospitalizations (blue line) plotted against weekly wastewater SARS-CoV-2 RNA copies per mL mean values (yellow line) from counties for which there were available data in a given week (Barnstable, Berkshire, Bristol, Essex, Franklin, Hampshire, Middlesex, Nantucket, Plymouth, Suffolk, Worcester), Massachusetts, February 2020 through June 2022 in Massachusetts. ^5^ Figure 1, Panel C is shown for comparison between documented cases and wastewater concentrations. Supplemental Figure 2C shows the cumulative Covid-19-associated deaths (blue line) plotted against weekly wastewater SARS-CoV-2 RNA copies per mL mean values (yellow line) from counties for which there were available data in a given week (Barnstable, Berkshire, Bristol, Essex, Franklin, Hampshire, Middlesex, Nantucket, Plymouth, Suffolk, Worcester), Massachusetts, February 2020 through June 2022 in Massachusetts.^5^ Supplemental Figure 2D shows the cumulative Covid-19-associated deaths (blue line) plotted against weekly confirmed new Covid-19 cases (yellow line), Massachusetts, February 2020 through June 2022 in Massachusetts.^5^

**Supplemental Figure 3.**
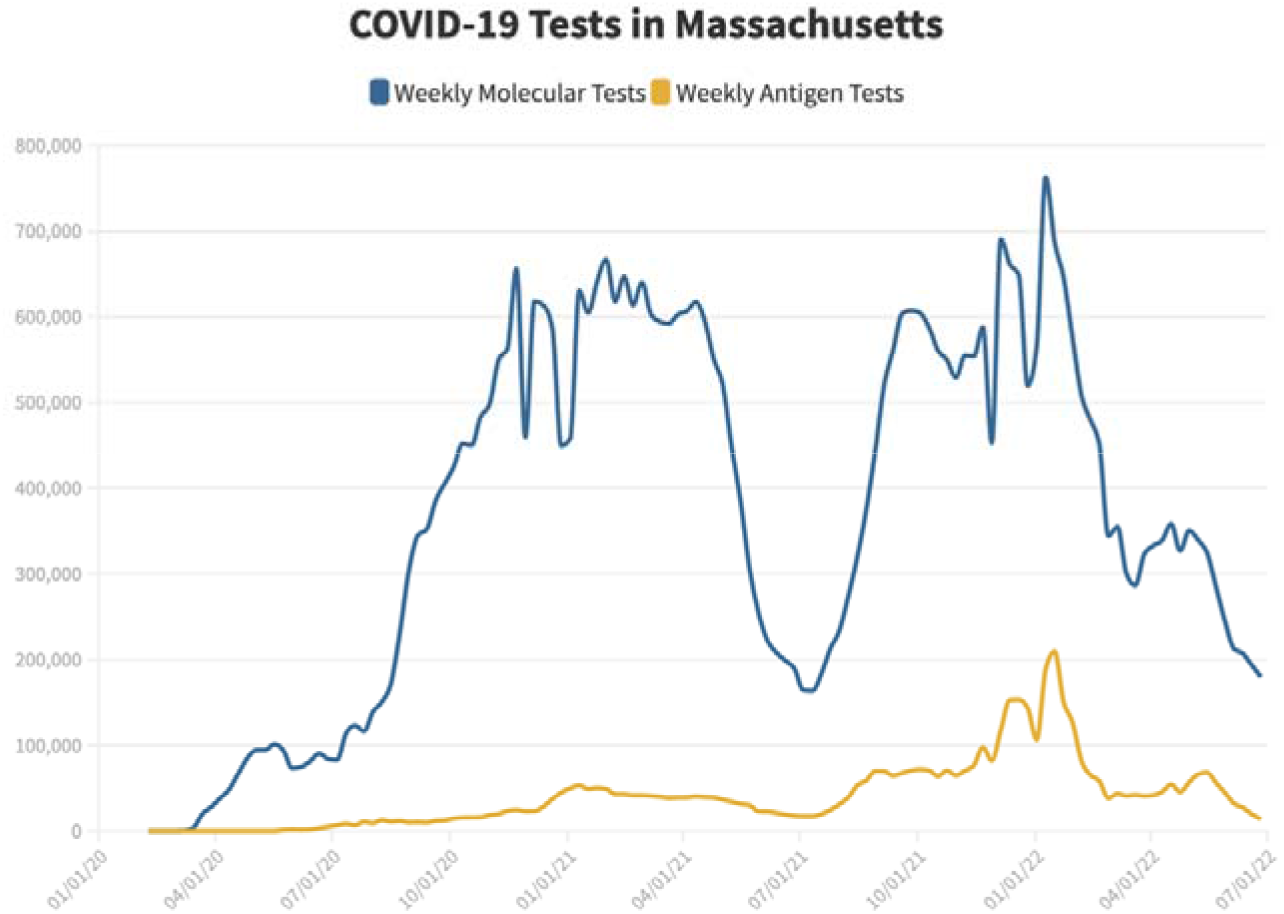
Weekly number of reported molecular test results (blue line) and antigen test results (yellow line), Massachusetts, August 8, 2020 through June 25, 2022.^5^

**Supplemental Figure 4a.**
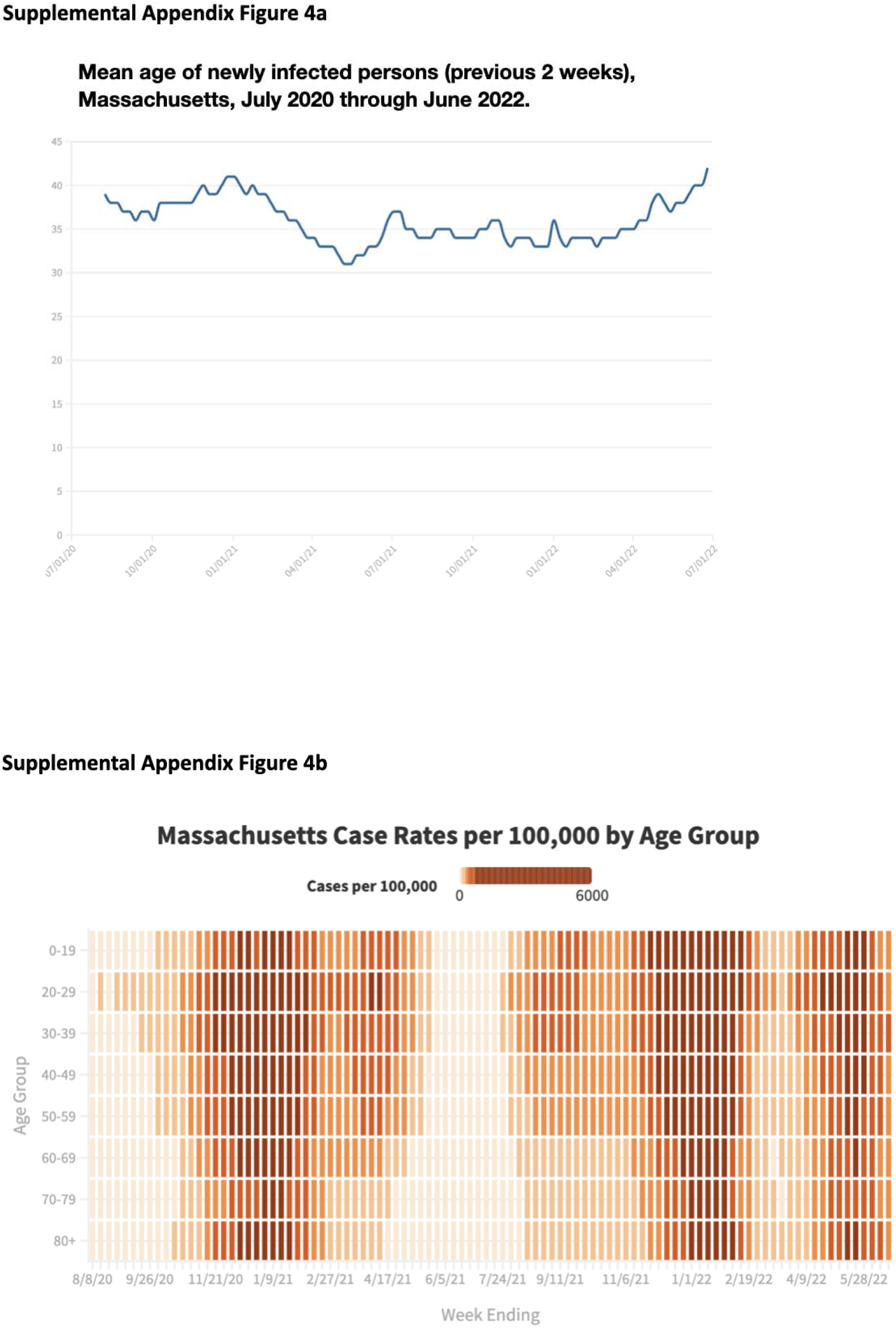
Weekly mean age of newly infected Covid-19 persons (previous two weeks), Massachusetts, March 2020 through June 2022.^5^ Supplemental Figure 4b. Weekly rates (per 100,000 residents) of newly infected Covid-19 persons by age group, Massachusetts, August 8, 2020 through June 18, 2022.^5^

